# The Effects of the Timing of Diuretic Administration and Other Covariates on Urine Volume in Hospitalized Patients

**DOI:** 10.1101/2022.12.27.22283947

**Authors:** Sara Abbaspour, Aaron D. Aguirre, M. Brandon Westover, Elizabeth B. Klerman

## Abstract

“What time should I take my medicine?” is an increasingly important question. Current knowledge of time-of-day effects for specific medications in hospitalized patients with cardiovascular disease is very limited. In such patients, increased medication efficiency could potentially reduce dose use and/or the length of time in the Intensive Care Unit (ICU) and/or hospital – potentially improving patient outcomes and patient and family quality of life, and reducing financial costs. We studied whether the time of day or night a patient is given a diuretic affects urine volume response. In this observational study, data were from 7,704 patients (63% male, 18 to 98 years old) admitted to one hospital’s acute care cardiac units, cardiac ICUs, cardiac surgery ICUs, and/or non-cardiac ICUs, who received intravenous furosemide (a diuretic), had measurements of urine volume, were hospitalized for ≥ 3 days between January 2016 to July 2021 and were older than 18 years. We used machine learning (ML) techniques to analyze the data. The ML technique identified factors that were expected to predict urine volume response to the diuretic: sex, age, medication dose and time, creatinine concentration, diagnosis, and hospital unit. The ML technique also identified medication administration time 00:00–06:00 as a predictor of higher urine volume response. Randomized controlled trials should be conducted to quantify the relative effect of modifiable factors, such as time of medication administration.

## Introduction

Circadian rhythms are physical, mental, and behavioral changes that display a period of approximately 24 h. These rhythms influence almost all areas of physiology, including the sleep-wake cycle, body temperature, blood pressure, and heart rate^1,2^. Healthy individuals produce more urine during the day than at night, and there are circadian and time-of-day (“diurnal”) rhythms in urinary sodium, potassium, and chloride excretion ^3–9^. Disruption of these rhythms is associated with hypertension and cardiovascular disease ^10^.

Chronomedicine aims to incorporate knowledge of biological rhythms to increase treatment effectiveness and decrease side effects ^1,11–13^;. Chronomedicine has demonstrated clinical benefits in hypertension, hypercholesterolemia, cardiac arrhythmias, and ischemic heart disease, cancer, diabetes, and other areas ^14–22^. The knowledge of medication time-of-day effects in hospitalized patients with cardiovascular disease is very limited. For hospitalized populations, there is tremendous potential benefit from any increased efficiency of medications. Another advantage of studying hospitalized patients is that there is a great source of accurately recorded data (e.g., medication dose and time, diet, frequently measured patients’ health outcomes such as vital signs or urine volume) to use for quantifying the impact of the timing of medication administration on patients’ health outcomes and the interactions of these variables. Adding a recommended time of day is a low-cost change (i.e., not requiring the development of a new medication or other intervention) that can be implemented almost immediately. Time-of-day differences in the efficacy of these interventions could be important for both understanding basic science and for potentially reducing doses given while achieving similar results.

In this study, we used machine learning (ML) methods to quantify the impact of the timing of diuretic medication administration and other covariates (e.g., sex, age, weight, medication dose, creatinine concentration, fluid intake, diagnosis, and hospital unit) on urine volume of hospitalized patients in Acute Cardiac Care and different Intensive Care Units (ICUs) The cause of the need to increase urine volume in patients in hospital ICUs)may be heart or renal failure (with fluid retention), fluids given during surgery, or other causes. Diuretics (e.g., furosemide) may be given to increase the urine flow rate; they have a rapid onset of response within the first few minutes after intravenous (IV) diuretic administration. ML is a powerful technique for diagnosis, detection, and prediction in medicine. Studies have used ML-based approaches to identify the most important clinical factors (e.g., sex, age, lab test results, temperature, and heart rate) in the prediction of volume responsiveness in patients with oliguric acute kidney injury in critical care^23^, identify the most critical factors in predicting the prevalence of stroke^24^, identify modifiable factors that influence COVID-19 vaccine side effects^25^ explain the contribution of different variables (e.g., age, tumor size, and the number of removed lymph nodes) in the prediction of 10-year overall survival of breast cancer^26^, and predict the risk of hypoxemia during general anesthesia and provide explanations of the risk factors (e.g., age, sex, BMI, blood pressure, temperature, and medication)^27^.

## Materials and Methods

### Data

The dataset was created from Mass General Brigham (MGB, Boston MA, USA) electronic health records (EHR). The inclusion criteria were everyone (i) admitted to Massachusetts General Hospital Acute Care Cardiac units, Cardiac ICUs, Cardiac Surgery ICUs, and Non-cardiac ICUs, (ii) who received IV furosemide, (iii) who had measurements of urine volume, (iv) who were hospitalized for ≥ 3 days between January 2016 and July 2021, and (v) were age ≥ 18 years. Patients differed in the reasons for furosemide administration (e.g., heart failure, post-surgery).

Variables considered for analysis were selected based on expert knowledge and their availability in EHR. Variables used were age, sex (female and male), weight, medication dose and time, lab test results (e.g., creatinine), fluid intake, medical condition/diagnosis (i.e., heart failure, acute kidney disease, chronic kidney disease, and cardiomyopathy), hospital unit, and urine volume and time. 0.8% of the data samples were removed because of missing values of creatinine (0.6%) and weight (0.2%). Weights > 200 kg or < 40 kg were not used (0.8% of the data points). B-Type Natriuretic Peptide Test was initially considered for analysis but there was not enough recording in EHR data for this variable (95% missing values); therefore, it was not included in the final feature set. The outcome of interest was urine volume rates in the hour after the time of medication administration normalized by the most recent (not older than 24 hours before the medication administration) body mass index (BMI).

The study was approved by The Mass General Brigham Institutional Review Boards (IRB).

### Pre-processing

The dataset was prepared by calculating the amount of administered medication rates by the time of day in hourly bins (sampling rate = one hour). The fluid intake rates and urine volume rates relative to medication administration in hourly bins were calculated. The calculations were performed using Resample function in Pandas package (version 1.3.4) in Python. Medication administrations were included in analyses when there were no other diuretic medications during two hours before through one hour after that dose, and no more than 4 hours gap between urine volume and fluid intake measurements. For each administered medication, urine volume rate normalized by BMI and fluid intake rate 2 hours before medication, urine volume rate normalized by BMI and fluid intake rate 1 hour before medication, and urine volume rates normalized by BMI in the hour after the time of medication administration were used for analysis. Time of day groupings were 00:00-05:59, 06:00-11:59, 12:00-17:59 or 18:00-23:59. Categorical variables (i.e., sex, time of day, diagnosis, and hospital units) were converted into dummy variables using OneHotEncoder, a scikit-learn (version 1.0.1) preprocessing package in Python. To avoid collinearity effect between the input variables, a Variance Inflation Factor analysis was conducted (threshold = 5) ^28^. BMI was calculated using the standard definition.

### Machine Learning Model

Machine Learning (ML) techniques use statistical methods to quantify relationships between input variables (e.g., patients age, sex, race, medical condition) and the target variables (e.g., health outcomes) in large datasets. Extreme Gradient Boosting (XGB) a tree-based machine learning model was selected because of its execution speed and performance ^26^, the advantage of high interpretability and the possibility of identifying the strongest predictors by applying a model explanation such as Tree Explainer ^28^. XGBRegressor from Xgboost package version 1.4.2 in Python (with max_depth = 4, number of estimators = 100, and learning rate = 0.1) was applied to the input variables to predict absolute the normalized urine volume rate in the hour after the time of medication administration. The model was parametrized using a randomized search of different parameter settings with a 5-fold cross validation.

### Evaluation

A stratified k-fold (k=5) cross-validation was used to validate the performance of the ML model. This method uses a large part of the data (80% of the data) to train the model, and a small part of the data (20% of the data) to test the model. No data from the same subject was used in the training and testing sets. Since some individuals were represented by more samples than others, a weight (1/number of samples per individual per hospital admit) was assigned to each data point in the dataset. The stratified cross-validation was repeated 10 times and the average and the standard deviation (SD) of mean absolute error (MAE) score (equation 1) was calculated. A baseline MAE was calculated by predicting the mean target value from the training dataset. A model with better MAE than the baseline MAE has prediction skill.

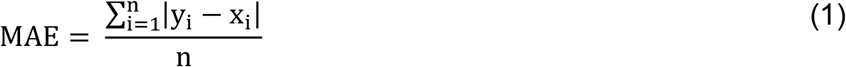

### Explainability

After the ML model is built and evaluated for its performance, model explanation techniques can be used to identify the most important predictors of urine volume rates and understand the contribution (magnitude and direction) of each of the input variables to the prediction of the urine volume rate values^26^. We chose SHapley Additive exPlanations (SHAP) values that are based on a game theory method for assigning an importance value to variables based on their contribution to a prediction ^28^. SHAP values were calculated using the trained ML model for each datapoint in the training dataset (i.e., the input values per medication administration). SHAP values were generated using the SHAP package (version 0.39.0 in Python). These values were used to obtain a visualization of the overall feature importance for the model. Then, SHAP boxplot (by applying a seaborn [version 0.11.2] boxplot package in Python) for categorical variables and SHAP scatter plot for continuous variables were generated to show how the features contributed to models’ output. All analyses were performed using open-source libraries in Python 3.7.

## Results

The data include 38,338 medication administrations in 7,704 patients during 8,324 hospital admissions (Table 1). Of these patients, 63% were male; ages ranged from 18 to 98 years old; weight ranged from 40 to 200 kilograms; medication dose ranged from 1 to 240 mg/hr; creatinine concentration ranged from 0.17 to 12 mg/dl. Of the hospital admits, 25% had admission diagnoses of heart failure, 6% acute kidney disease, 4% chronic kidney disease, 24% cardiomyopathy, and 41% had other reasons for hospital admit. 7% were admitted to Acute Care Cardiac Unitss, 15% to Cardiac ICUs, 37% to Cardiac Surgery ICUs, and 41% to Non-cardiac ICUs. 16% of medication administrations were from 00:00 to 05:59, 33% from 06:00 to 11:59, 33% from 12:00 to 17:59, and 18% from 18:00 to 23:59.

**Table 1.** Number of individuals and (%) percent of total patients; Number of medication administrations and (%) percent of total medication administrations. Note, due to rounding, not all all percentages sum to 100%

| **Variables** | **Number of individuals**<br>**N(%)** | **Number of medication administration**<br>**N(%)** |
| --- | --- | --- |
| **Sex** |  |  |
| Female | 2,876 (37) | 14,856 (39) |
| Male | 4,828 (63) | 23,482 (61) |
| **Total** | **7,704 (100)** | **38,338 (100)** |
| **Admit Diagnosis** |  |  |
| Heart Failure | 1,855 (24) | 8,696 (23) |
| Acute Kidney Disease | 481 (6) | 2527 (7) |
| Chronic Kidney Disease | 340 (4) | 1751 (5) |
| Cardiomyopathy | 1,789 (23) | 8,520 (22) |
| Other reasons for visit | 3,239 (42) | 16,844 (44) |
| **Total** | **7,704 (100)** | **38,338 (100)** |
| **Hospital Unit** |  |  |
| Acute Care Cardiac Unit |  | 2,767 (7) |
| Cardiac ICU |  | 5,649 (15) |
| Cardiac Surgery ICU |  | 14,300 (37) |
| Non-cardiac ICU |  | 15,622 (41) |
| **Total** |  | **38,338 (100)** |
| **Medication Time** |  |  |
| 00-06H |  | 2,767 (16) |
| 06-12H |  | 5,649 (33) |
| 12-18H |  | 14,300 (33) |
| 18-00H |  | 15,622 (18) |
| **Total** |  | **38,338 (100)** |

Ten features (sex, age, weight, medication dose, creatinine, fluid intake, admit diagnosis, hospital unit, time of day of medication administration, urine volume and fluid intake 2 hours before medication, and urine volume and fluid intake 1 hour before medication) were used to build a predictive ML model. The model showed predictive MAE score of 4.1 (SD = ± 0.03) vs 5.4 baseline MAE for normalized urine volume rates.

A SHAP feature importance plot was created using mean absolute SHAP values of the ML model for predicting absolute urine volume rates normalized by BMI (Figure 1); this plot orders the input variables (top to bottom along the y-axis) according to their importance to the ML model. The five most important predictors to predict absolute urine volume rates were, in descending order: urine volume the hour before medication administration, creatinine, weight, sex, and age (Figure 1).

**Figure 1.**
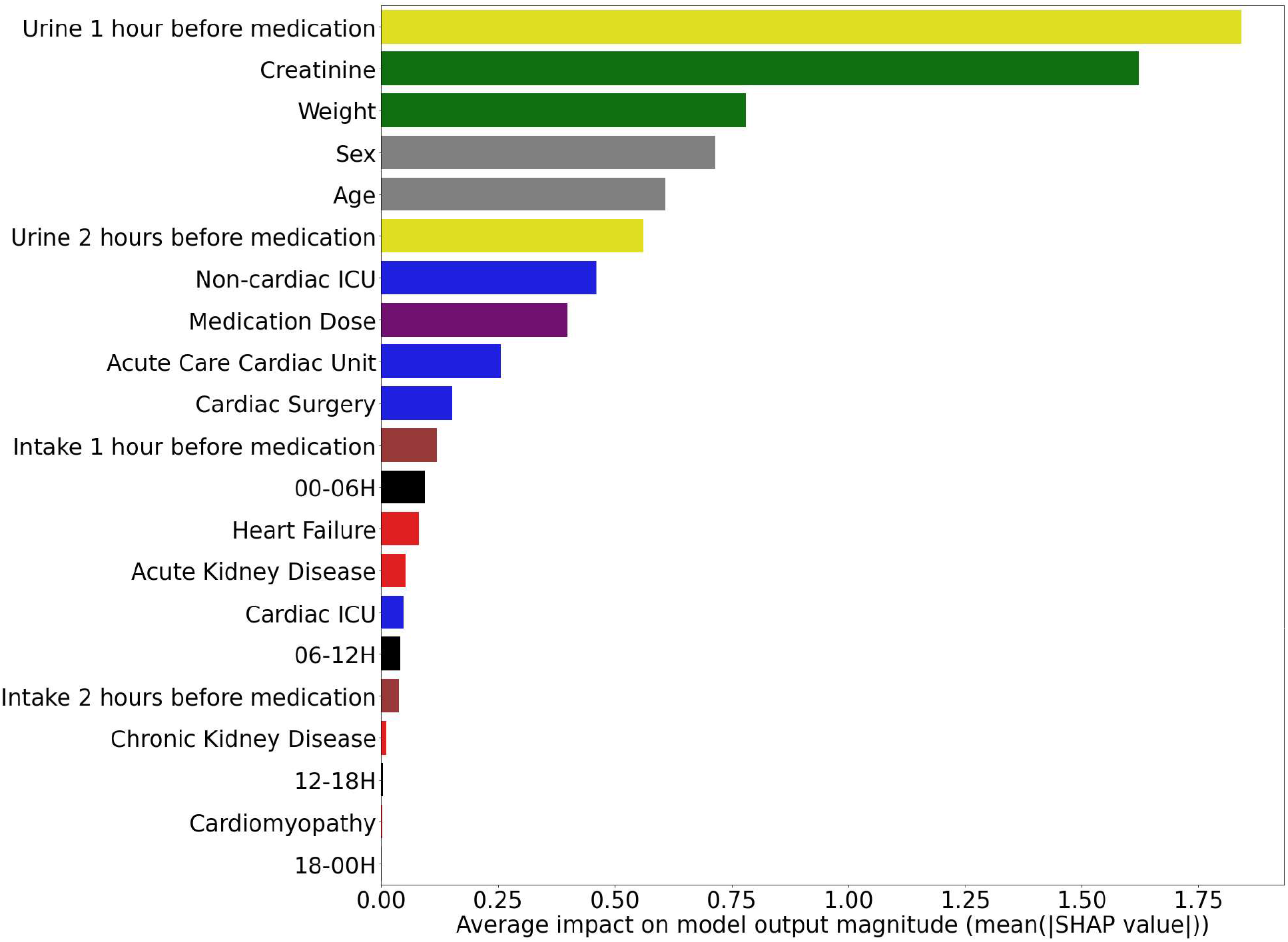
Feature importance plot using the mean absolute SHAP values.

The SHAP boxplot for binary variables (Figure 2) shows the direction of impact of the categorical variables on the model output in predicting absolute urine volume rates. Positive SHAP values are associated with higher urine volume rate value predictions and negative SHAP values are associated with lower urine volume rate value predictions. Male, being admitted to Non-cardiac ICU and medication administration between 00:00 to 05:59 were associated with more urine volume output in response to a diuretic (Figure 2). Female, diagnosis of heart failure, acute kidney disease, and chronic kidney disease, and being admitted to the Acute Care Cardiac Unit and Cardiac Surgery ICU were associated with less urine volume in response to a diuretic (Figure 2).

**Figure 2.**
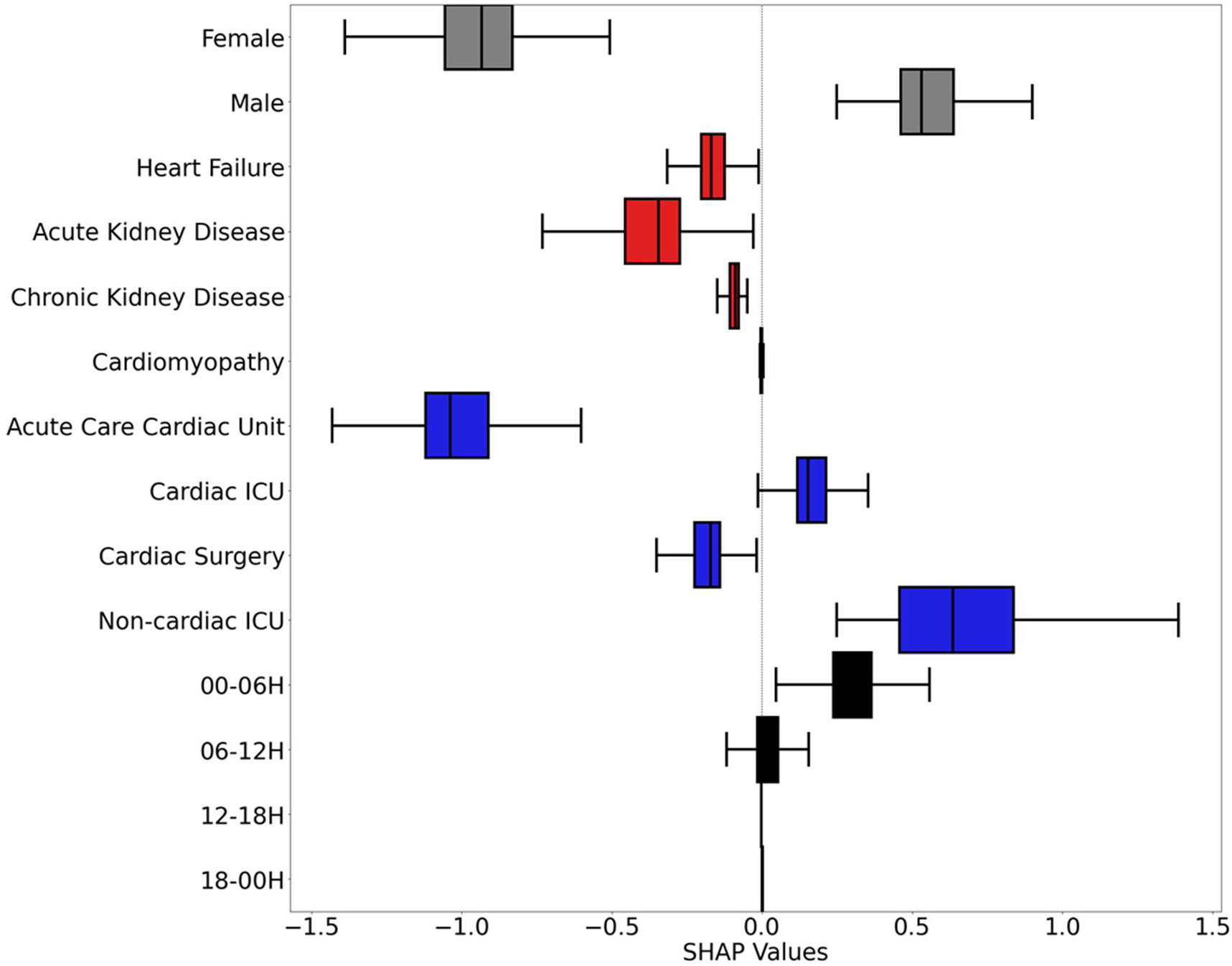
SHAP value boxplot that shows the direction of impact of each binary variable on model’s output. Positive SHAP values are indicative of a higher urine volume value prediction, while negative SHAP values are indicative of a lower urine volume value prediction; vertical lines indicate 5th and 95th percentiles. Box limits indicate 25th and 75th percentile and vertical line within the box indicates 50th percentile.

For the continuous variables, younger adults, a medication dose between 25 to 75 mg/hr, and lower creatinine concentrations were associated with more urine volume in response to a diuretic (Figure 3 and 4). There was a high association between higher creatinine concentration and higher dose of medication (Figure 4).

**Figure 3.**
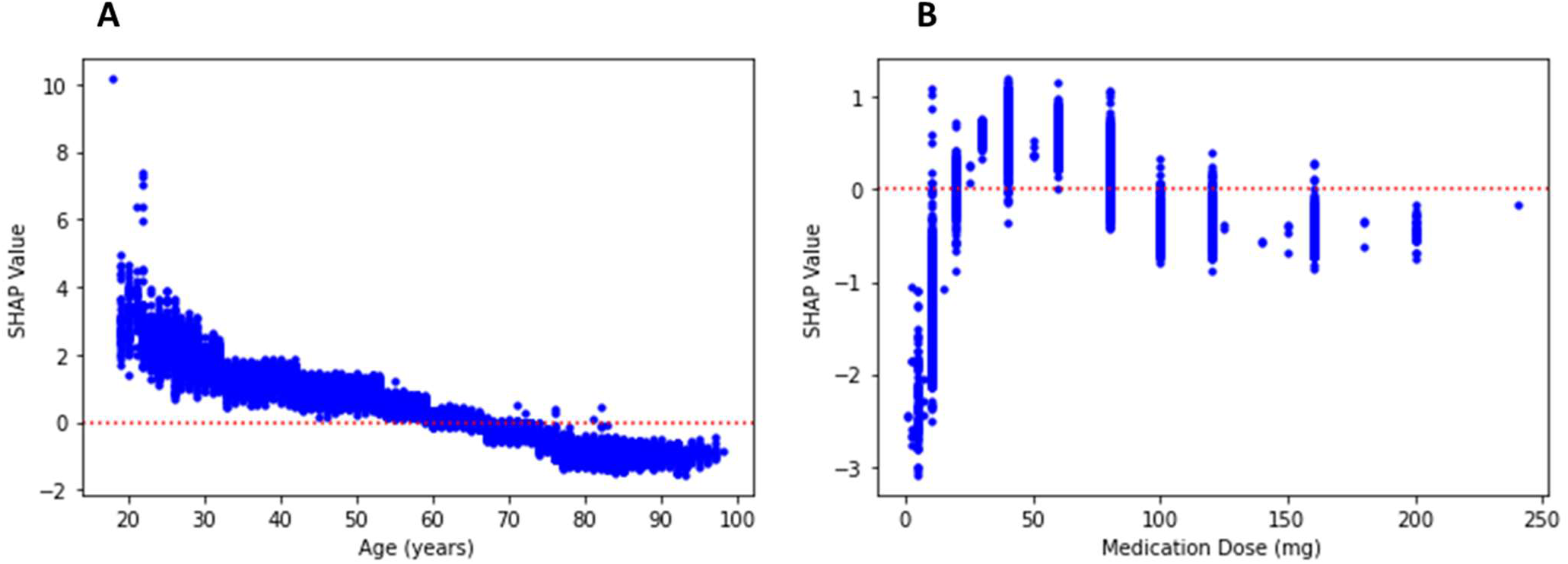
SHAP scatter plot for continuous variables A: age, B: medication dose and C: creatinine concentration showing the impact of these variables on model’s output.

**Figure 4.**
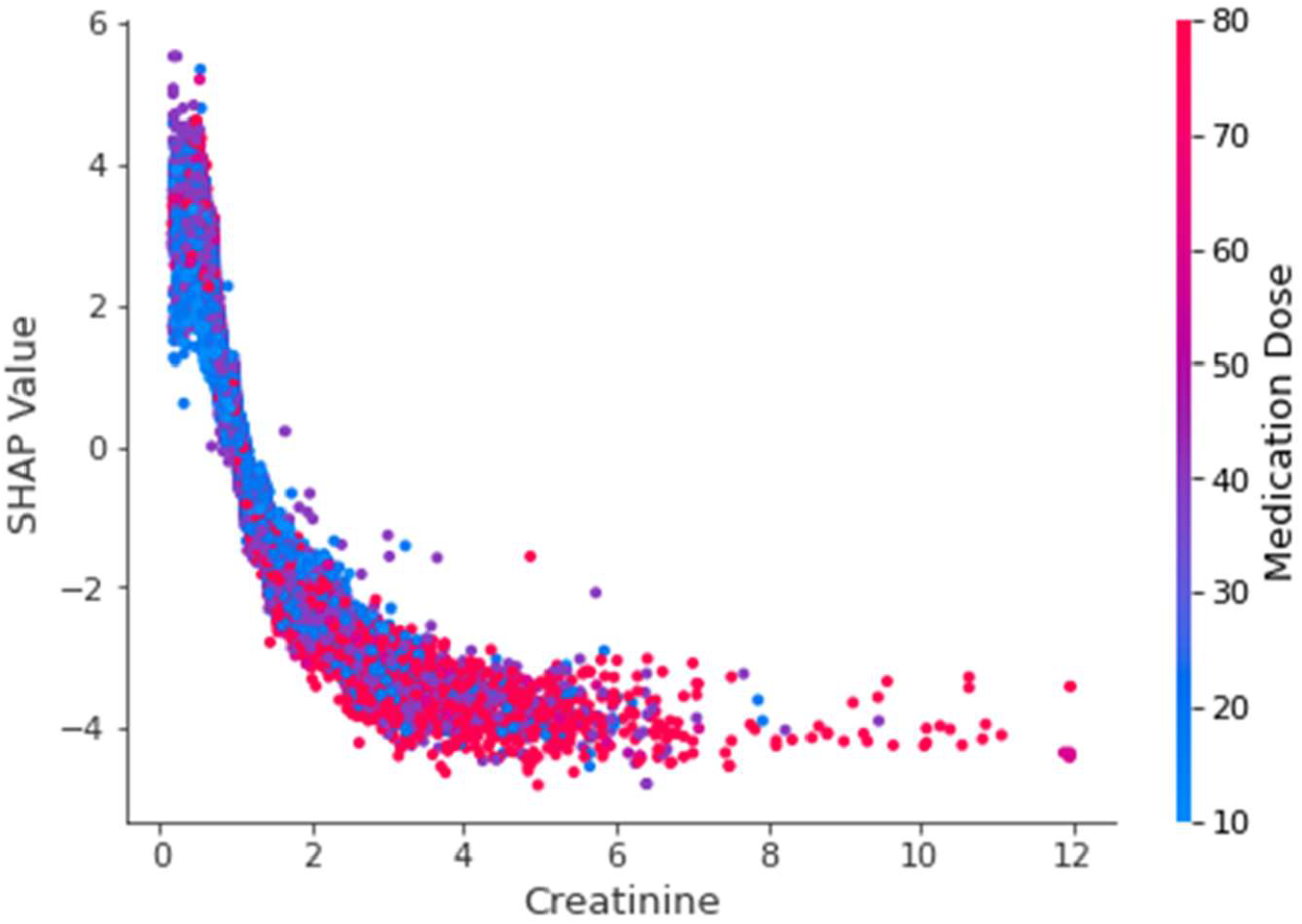
SHAP scatter plot for creatinine concentration showing the impact of this variable on model’s output (red: higher dose of medication and blue: lower dose of medication).

## Discussion

We used an explainable ML method to quantify the effect of time of day of a diuretic administration and other covariates on urine volume in hospitalized patients. Our results are consistent with several other publications that documented relationships between urine volume in response to a diuretic and sex, age, diagnosis of heart or kidney disease, and creatinine levels. Our novel findings are of time-of-day effects of IV furosemide on urine volume, with a significantly larger effect of medication administration for doses between midnight and 6am in a ML model that included fluid intake as a variable. Future work will include time-of-day variation in urine volume rates when no diuretic has been given.

Chronomedicine aims to incorporate knowledge of biological rhythms in clinical, including preventive, care ^29^. Health care professionals are beginning to consider time-of-day in their diagnosis and treatment administration. The information about time-of-day may be used to better define relevant physiology and improve clinical care in outpatient and inpatient populations. The availability of data from EHR including telemetry will be important^30–32^. For example, altering the time of an intervention to increase efficacy would be a relatively low-cost and scalable change in practice.

EHR provide the opportunity to improve healthcare. Handling these large and complex datasets requires special computational techniques that can deal with these datasets. ML techniques have broad applications in healthcare and are helpful in identifying patterns in large datasets^33^. Developments in the area of ML and model explanation, and strong methods to compute and visualize the magnitude and direction of impact of input variables on model’s outputs, can help translate knowledge from science to practice ^28,34^. Given our multidimensional datasets, the application of ML can be useful since its strength includes dealing with many input variables.

Limitations of this work are that the data are from an observational study. Randomized clinical trials should be performed to further test our hypotheses of time-of-day influences on immediate drug effects, longer-term effects on multiple metrics of patient health and quality of life, length of stay in ICUs and in-hospital total, and financial costs. Basic science studies should also be done to better define physiology and develop new treatments. Education for health care providers (e.g., physicians, nurses, and pharmacists) about time-of-day effects and variation in clinical metrics can also be implemented.

## Conclusion

In this study, we used XGB, a ML model to predict the urine volume rates in the hour after the time of medication administration using multiple potentially relevant variables (e.g., sex, age, medication dose and time, creatinine, fluid intake, admit diagnosis, and hospital units). We then used a model explanation technique (SHAP) to identify the important predictors of urine volume rates and explain the effect of the input variables on model’s output. Our results demonstrate that age, sex, medication dose, creatinine concentration, admit diagnosis, hospital units, and time of day of medication administration (00:00– 06:00) are associated with a significantly higher predicted urine volume response to a diuretic. Time-of-day differences in the efficacy of interventions could be important for both understanding basic science and for potentially reducing doses given while achieving similar results. Adding a recommended time of day is a low-cost change (i.e., not requiring the development of a new medication or other intervention) that can be implemented almost immediately.

## Data Availability

All data produced in the present study are available upon reasonable request to the authors

## Notes

**Support:** MBW: NIH R01-NS102190, R01-NS102574, R01-NS107291, RF1-AG064312, RF1-NS120947, R01-AG073410, R01-HL161253 and NSF 201443). EBK: NIH R01-NS099055, U01-NS114001, U54-AG062322, R21-DA052861, R21-DA052861, R01-NS114526-02S1, R01-107064; DoD W81XWH201076; and Leducq Foundation for Cardiovascular Research

**Relevant Conflicts of Interest** SA, ADA, MDW: none. EBK: Consulting: American Academy of Sleep Medicine Foundation, Circadian Therapeutics, National Sleep Foundation, Sleep Research Society Foundation, Yale University Press. Travel: European Biological Rhythms Society. Other: Partner owns Chronsulting.

### Competing Interest Statement

SA, ADA, MDW: none.
EBK: Consulting: American Academy of Sleep Medicine Foundation, Circadian Therapeutics, National Sleep Foundation, Sleep Research Society Foundation, Yale University Press. Travel: European Biological Rhythms Society. Other: Partner owns Chronsulting.

### Funding Statement

MBW: NIH R01-NS102190, R01-NS102574, R01-NS107291, RF1-AG064312, RF1-NS120947, R01-AG073410, R01-HL161253 and NSF 201443).
EBK: NIH R01-NS099055, U01-NS114001, U54-AG062322, R21-DA052861, R21-DA052861, R01-NS114526-02S1, R01-107064; DoD W81XWH201076; and Leducq Foundation for Cardiovascular Research

### Author Declarations

The IRB of The Mass General Brigham gave ethical approval for this work

